# Investigating psychobiological causes and mechanisms in functional seizures and functional motor symptoms: Study protocol

**DOI:** 10.1101/2024.05.24.24307863

**Authors:** Susannah Pick, Anthony S. David, Mark J. Edwards, Laura H. Goldstein, John Hodsoll, L. S. Merritt Millman, Timothy R. Nicholson, A.A.T.S. Reinders, Biba Stanton, Joel S. Winston, Mitul A. Mehta, Trudie Chalder, Matthew Hotopf

## Abstract

**Introduction:** Advances have been made in understanding the aetiology of functional neurological disorder (FND); however, its pathophysiological mechanisms have not been definitively demonstrated. Evidence suggests interacting roles for altered emotional processing and interoception, elevated autonomic arousal, and dissociation, but there is limited evidence demonstrating their causal influence on specific FND symptoms. Our superordinate aim is to elucidate potentially shared and distinct aetiological factors and mechanisms in two common FND subtypes, functional seizures (FS) and functional motor symptoms (FMS).

**Methods:** This study has a multimodal, mixed between- and within-groups design. The target sample is 50 individuals with FS, 50 with FMS, 50 clinical controls (anxiety/depression), and 50 healthy controls. Potential aetiological factors (e.g., adverse life events, physical/mental health symptoms, dissociative tendencies, interoceptive insight/sensibility) will be assessed with a detailed medical history interview and self-report questionnaires. A laboratory session will include a neurocognitive battery, psychophysiological testing, cardiac interoception and time estimation tasks and an isometric handgrip task. A subsample will undergo magnetic resonance imaging, including structural, resting-state and task-based scans combined with psychophysiological recording. Remote monitoring with ecological momentary assessment and wearables will measure variability in FND symptoms and in patients’ daily lives and their potential predictors/correlates for ≥2 weeks. Longitudinal follow-ups at 3, 6, and 12-months will monitor longer-term outcomes in the clinical groups.

**Discussion:** This study employs multimodal research methods to rigorously examine several putative mechanisms in FND, at subjective/experiential, behavioural, and physiological levels. The study will test causal hypotheses about the role of altered emotional processing, autonomic arousal, dissociation and interoception in the initiation or exacerbation of FND symptoms, directly comparing these processes in FS and FMS to healthy and clinical controls. This is the first study of its kind, with potential to reveal important targets for prevention and treatment of FND in future.

## Introduction

Functional neurological disorder (FND) is a complex neuropsychiatric disorder defined by the presence of neurological symptoms (motor, sensory, seizure) that are not caused by or compatible with identifiable neuropathology (APA, 2013; WHO, 2019). FND is commonly associated with impaired quality of life, diminished functioning, psychological distress and significant healthcare costs (Carson & Lehn, 2016). Individuals with FND experience considerable stigma and barriers to accessing appropriate treatment, which is exacerbated by uncertainty regarding causation and mechanisms.

It is now accepted that there is a wide range of psychological, social and biological aetiological factors associated with FND, including stressful life events, mental health symptoms/disorders, relationship problems and significant physical injuries and illnesses (Brown & Reuber, 2016; Morsy et al., 2021; Reuber, 2009). However, precisely how these factors contribute to FND is still not fully understood and there is a need for more rigorous research into the mechanisms underlying FND symptoms.

Recent perspectives have highlighted the potential role of altered emotional information processing and elevated autonomic arousal in FND (Brown & Reuber, 2016; Drane et al., 2020; Pick et al., 2019). Previous experimental evidence has shown that people with FND exhibit differences in autonomic and/or subjective responses to affective stimuli (e.g., Pick, Mellers & Goldstein, 2018a; Pick et al., 2023; Roberts et al., 2012), reduced recognition and attentional biases to emotional facial expressions (Bakvis et al., 2009; Pick, Mellers & Goldstein, 2016; 2018b), altered bodily awareness (interoception) (Koreki et al., 2020; Pick et al., 2020; Williams et al. 2021), and divergent patterns of neural activation during exposure to affective stimuli (e.g., Aybek et al., 2015; Espay et al., 2018). There is also accumulating evidence to support the view that dissociation may be an important underlying pathophysiological process (e.g., Campbell et al., 2022; Koreki et al., 2020; Pick, Mellers & Goldstein, 2017; Pick et al., 2020).

Despite advances in the understanding of FND, there remain some important unanswered questions. Firstly, few studies have examined explicitly the potential temporal relationships between key pathophysiological processes (e.g., autonomic arousal, limbic hyperactivation) and the occurrence of FND symptoms (Pick et al., 2019). Evidence for causal relationships between putative mechanistic processes and FND symptoms is therefore limited, with many previous studies demonstrating only correlational relationships. There is a paucity of studies directly comparing potential causal factors and pathophysiological mechanisms in different FND subtypes, and the possible links between background aetiological factors and mechanistic processes in these subgroups require further examination. Furthermore, there is little evidence linking specific aetiological factors and mechanistic processes to longer-term outcomes in FND.

### Aims & objectives

The overall aim of this project is to elucidate the shared and distinct psychobiological causes and mechanisms of functional seizures (FS) and functional motor symptoms (FMS), using a novel combination of multi-modal research methods.

The primary objective is to examine the hypothesised influence of autonomic arousal and limbic system hyperactivation on subjective FS and FMS symptoms (Pick et al., 2019).

Secondary objectives are as follows:

- Assess a range of biological, psychological and social aetiological factors in people with FS and FMS, including adverse life events, psychological distress and dissociative tendencies.
- Examine relationships between background factors (e.g., adverse experiences), psychobiological mechanisms (e.g., autonomic arousal, limbic hyperactivity, bodily awareness) and clinical outcomes (i.e., symptom severity, functioning, quality of life).
- Identify the similarities and differences in background factors and pathophysiological processes in the FND groups (FS, FMS) relative to individuals with elevated psychological symptoms (anxiety, depression), who do not experience neurological symptoms.
- Examine factors that might trigger the occurrence or worsening of FS and FMS symptoms in patients’ daily lives in real-world contexts, including the possible influence of autonomic arousal, sleep disturbance, physical exertion, daily events and mood variations.

To achieve these objectives, the following methods are being employed:

- An in-depth interview and self-report questionnaires to capture data on a range of possible predisposing, precipitating and perpetuating factors.
- Standardised neurocognitive tests to examine core cognitive domains implicated in the pathophysiology of FND, including attention (Edwards et al., 2012), executive functions (Brown & Reuber, 2016), and social cognition (Pick et al., 2019).
- Experimental and psychophysiological measures to probe behavioural, cognitive and physiological responses to bodily sensations and affectively significant stimuli.
- Neuroimaging to assess structural and functional brain differences in people with FS and FMS, compared to both healthy and clinical controls.
- Remote monitoring with smartphone applications and a wearable device to collect data on the possible antecedents of FS and FMS in ‘real-time’ and ‘real-world’ contexts.
- Remote follow-up sessions at 3, 6, and 12-months to examine longer-term clinical outcomes.

The primary hypotheses being tested are:

1. Individuals with FS and FMS will exhibit elevated autonomic activation (heart rate, electrodermal activity) in response to affectively significant stimuli and events, compared to healthy and clinical controls.
2. Elevated autonomic arousal and associated hyperactivation in limbic brain regions will temporally precede the occurrence or exacerbation of FS and FMS.

The secondary hypotheses include the following:

1. People with FS and FMS will display alterations in bodily awareness (interoception) and associated neural activity, compared to controls.
2. Participants with FS and FMS will exhibit differences in executive functioning (e.g., attentional allocation, response inhibition) and social cognition compared to controls, despite intact general cognitive functioning.
3. FS and FMS symptom severity will be exacerbated in daily life by emotionally salient events, dissociation, and negative affect.
4. Worse clinical outcomes (symptom severity, quality of life, functioning, psychological distress) will be predicted by adverse life event burden, elevated autonomic arousal, limbic hyperactivation, and greater psychological dissociation.

## Materials and methods

This study underwent peer-review by four independent reviewers during funding acquisition (Medical Research Council UK) and has been reviewed/approved by the Health Research Authority and an NHS Research Ethics Committee (‘Ethical considerations’). This is a single centre study; all research activities are taking place at, or being coordinated from, the Institute of Psychiatry, Psychology and Neuroscience, King’s College London.

### Study Design

This project has a mixed between- and within-groups design, including observational, experimental, cross-sectional and longitudinal measures. The FS and FMS samples are being compared to healthy and clinical control groups.

#### Primary endpoints

- FS and FMS occurrence and severity, measured with repeated momentary assessments during the laboratory, neuroimaging and remote monitoring procedures.
- Autonomic arousal, measured with heartrate and/or electrodermal activity levels during the laboratory, neuroimaging and remote monitoring procedures.
- Limbic and paralimbic system hyperactivation, measured with region-of-interest neuroimaging analyses.

#### Secondary endpoints

- FS and FMS severity, occurrence, and impact, measured with self-report questionnaires at baseline and 3, 6 and 12-month follow-up.
- Psychological dissociation, measured with momentary assessments during the laboratory, neuroimaging session and remote monitoring phased, and with validated scales at baseline and follow-up.
- Interoceptive accuracy and awareness, and associated functional brain activity (i.e., insula), measured with experimental tasks and self-report measures at baseline, during the laboratory session and at follow-up, and with region-of-interest analyses in the neuroimaging study.
- Cognitive functioning, measured with standardised neurocognitive tests and experimental tasks during the laboratory session.
- Work, social and physical functioning, measured with validated scales at baseline and at follow-up.
- Health-related quality of life, also measured with a validated questionnaire at baseline and follow-up.

### Pilot study

The design of this project has been informed by a pilot study which was conducted between July-October 2022, including 17 participants with FMS and/or FS and 17 healthy controls. The pilot study included all procedures proposed here, except the neuroimaging session. The aim was to test the acceptability and feasibility of the procedures, and to ensure that the tasks planned for the neuroimaging session were well-tolerated and valid. The 34 participants who entered the study completed all elements of the research. Feedback was elicited from participants, which has now been incorporated into this protocol. The findings of the pilot study have been reported elsewhere (Millman et al., 2023, 2024; Pick et al., 2023a,b).

### Patient/public involvement

An FND Patient and Carer Advisory Panel (FND-PCAP) was convened in 2022 to consult with the team throughout the project on the design, implementation and dissemination of the project. The FND-PCAP includes patients with FND, carers, and representatives of patient support organisations (FND Hope UK, FND Action).

### Participants

Fifty participants diagnosed with FS and 50 with FMS will be recruited in total, in addition to 50 healthy control participants and 50 matched clinical control participants. The groups will be frequency-matched for age, sex/gender, and handedness. The eligibility criteria are detailed below.

#### Inclusion criteria

All participants:

- 18-65 years old
- Normal or corrected eyesight
- Fluency in English language Participants with FS:
- A primary diagnosis of functional seizures made by a Consultant Neurologist/Epileptologist, in the absence of functional motor symptoms
- Currently meets DSM-5 criteria for FND
- Minimum seizure frequency of 2 per month, with premonitory symptoms Participants with FMS:
- A primary diagnosis of functional motor symptoms made by a Consultant Neurologist, in the absence of functional seizures
- Currently meets DSM-5 criteria for FND

Clinical control participants:

- Presence of anxiety disorder or major depression, confirmed with DSM-5 criteria during the screening interview (Quick SCID)

#### Exclusion criteria

All participants:

- Diagnosis of major comorbid cardiovascular (e.g., heart disease), active severe psychiatric disturbance (e.g., psychosis, alcohol, or substance dependence) or neurological disorder (e.g., epilepsy, multiple sclerosis) that would either confound the findings or impair the participant’s ability to participate
- Physical symptoms / disability impairing ability to perform tasks (e.g., severe/constant tremor, bilateral upper limb paralysis, daily seizures)
- Current participation in another interventional study (e.g., treatment trial, experimental study)

Neuroimaging study:

- Ineligibility to undergo MRI imaging, for example the presence of a cardiac pacemaker or other electronic device or ferromagnetic metal foreign bodies.
- Participant weight in excess of 126kg or physical dimensions such that the participant may not fit in the MRI scanner.
- A history of claustrophobia or participant reports symptoms triggered in confined spaces or participant feels unable to lie in an MRI scanner for a period of up to 90 minutes.
- Any other reason that in the opinion of the investigator may impact the safety of participants or the integrity of the data.

Healthy control participants:

- Diagnosis of FND
- Active major physical or mental health disorder Clinical control participants:
- Diagnosis of FND
- Active suicidality or non-suicidal self-injury

### Study procedures

Figure 1 details the study procedures including the timepoint at which each measure is administered.

**Figure 1.**
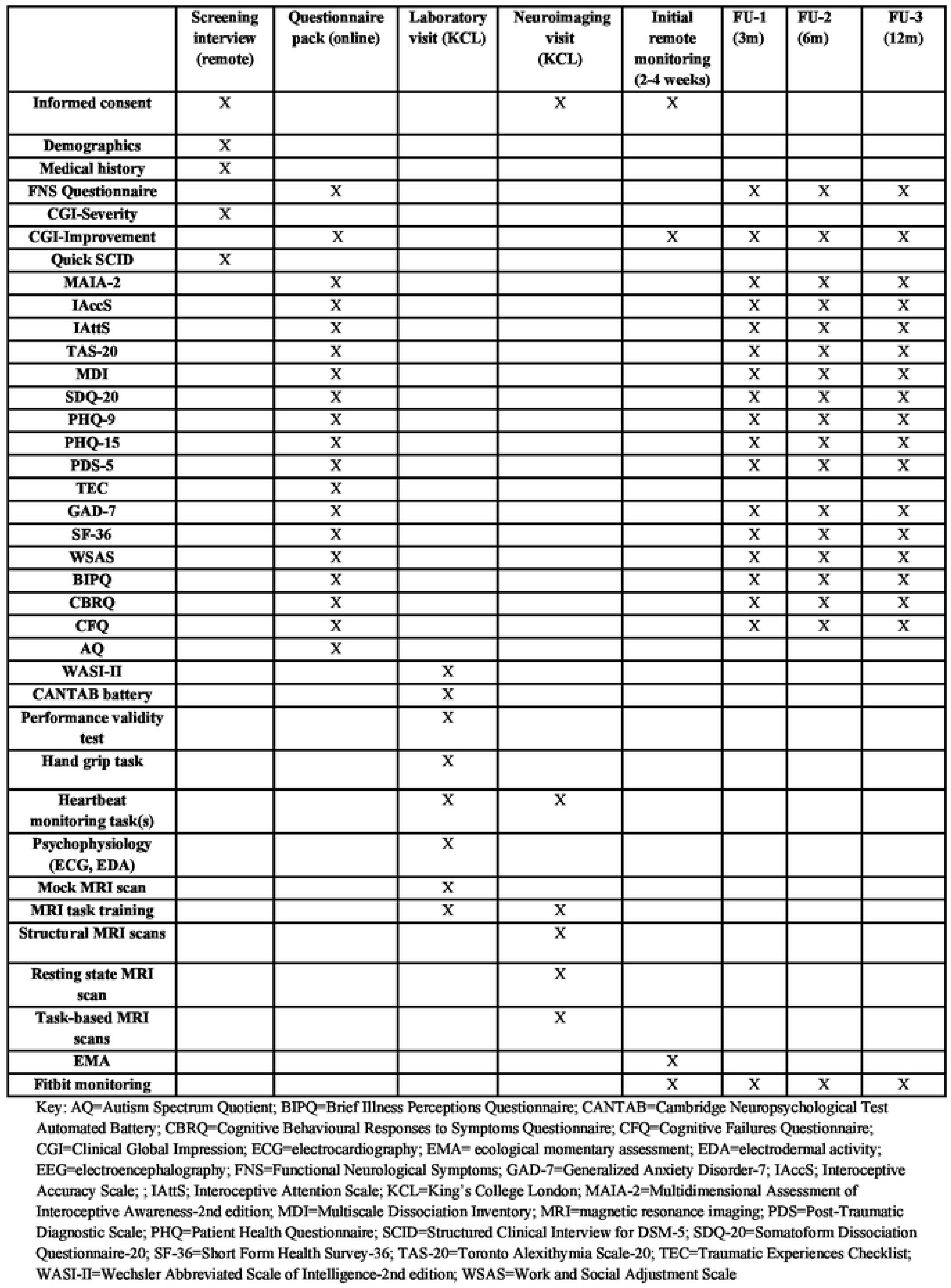
Flowchart of study procedures.

#### Participant recruitment

Recruitment of participants commenced in November 2023 and will end in December 2025 or when the target sample size is achieved, if earlier.

Participants with FND are being referred from neurology and/or neuropsychiatry services at the South London and Maudsley, King’s College Hospital, University College Hospitals and St George’s University Hospitals NHS Foundation Trusts. Participants with FND are also being recruited online, with advertisements circulated by charitable patient support websites (e.g., FND Hope UK, FND Action). Healthy and clinical control participants are being recruited from the community via advertisements in public places and social media platforms. Clinical control participants will also be recruited from existing KCL research cohorts (e.g., the Genetic Links to Anxiety and Depression study [https://gladstudy.org.uk/]).

Potential participants who express interest in the study and appear to be eligible at initial screening are provided with a Participant Information Sheet (PIS) and Informed Consent Form (ICF) by their clinician or the research team.

#### Informed Consent Procedures

There are three written informed consent procedures during this study, as follows:

1. Main consent form: Participants who appear to be eligible at initial contact are asked to complete a consent form electronically for completion of the screening interview, background questionnaires, and laboratory session. Consent for this stage is obtained at least 48 hours after participants received the PIS/ICF, unless a participant decides to participate sooner, in which case they are able to contact the research team directly.
2. Neuroimaging: Another ICF is completed for the MRI session, either electronically or in person. If seemingly eligible at initial screening, participants are given a separate neuroimaging PIS and ICF for consideration for at least 48 hours.
3. Remote monitoring: Participants complete another ICF, electronically or in person, for participation in the remote monitoring phase, prior to installing the smartphone applications on their mobile devices.

Consent is obtained at all timepoints by a member of the immediate research team, only after participants’ questions have been answered and a full, detailed explanation of the research activities has been provided.

#### Screening Procedures

All participants are screened initially for eligibility by either a member of the research team or a local clinician. Potential participants are assessed for eligibility briefly on the basis of current physical or mental health diagnoses, FND symptoms (if applicable), and medications. Those candidates appearing to meet the eligibility criteria are asked to provide informed consent prior to participating in a detailed screening interview with a senior member of the research team by telephone or online. During the interview, participants complete a detailed background sociodemographic and medical history questionnaire, followed by a brief structured clinical interview (Quick SCID; First & Williams, 2021). Eligibility for all elements of the study, including neuroimaging, are evaluated at this stage. Following the interview, participants are informed of the outcome. Should the candidate not be eligible to progress into the main study, they are given a detailed explanation and reimbursed for their time with a £10 shopping voucher.

### Schedule of Assessments

#### Online self-report questionnaires

Eligible participants are emailed a weblink to a set of self-report questionnaires (Qualtrics), which assess the following aetiological factors:

- Life events (Traumatic Experiences Checklist; Nijenhuis, Van der Hart, & Kruger, 2002)
- Trauma-related distress (Posttraumatic Stress Diagnostic Scale – Self-Report Version for DSM-5; Foa et al., 2016)
- Work and social functioning (Work and Social Adjustment Scale; Mundt, Marks, Shear, & Greist, 2002)
- Health-related quality of life (Short Form Health Survey – 36 item; Hays, Sherbourne, & Mazel, 1993)
- Psychological distress (Patient Health Questionnaire-9; Kroenke, Spitzer, & Williams, 2001 / Generalized Anxiety Disorder-7; Spitzer, Kroenke, Williams, & Lowe, 2006)
- Physical symptom burden (Patient Health Questionnaire-15; Kroenke, Spitzer, & Williams, 2002)
- Interoceptive abilities (Interoceptive Accuracy Scale, Murphy et al., 2020; Interoceptive Attention Scale, Gabriele et al., 2022; Multidimensional Assessment of Interoceptive Awareness-2, Mehling et al., 2018)
- Dissociative tendencies (Somatoform Dissociation Questionnaire-20; Nijenhuis, Spinhoven, Van Dyck, Van der Hart, & Vanderlinden, 1996; Multiscale Dissociation Inventory; Briere, 2005)
- Alexithymia (Toronto Alexithymia Scale-20; Bagby, Taylor, & Parker, 1994)
- Autistic traits (Autism Spectrum Quotient; Baron-Cohen, Wheelwright, Skinner, Martin, & Clubley, 2001)
- FND symptoms (bespoke questionnaire assessing the presence and severity of FND symptoms)
- Illness-related cognitions (Brief Illness Perceptions Questionnaire; Broadbent, Petrie, Main, & Weinman, 2006 / Cognitive Behavioural Responses Questionnaire; Ryan, Vitoratou, Goldsmith, & Chalder, 2018)
- Subjective cognitive symptoms (Cognitive Failure Questionnaire; Broadbent, Cooper, FitzGerald, & Parkes, 1982)

#### Laboratory session

Participants are next invited to attend a laboratory session, consisting of two parts separated by a break of approximately 30-60 minutes.

Part 1: Standardised neurocognitive testing

1. Wechsler Abbreviated Scale of Intelligence (second edition, WASI-II, Wechsler, 2011) assesses general cognitive abilities.
2. A CANTAB Connect cognitive test battery (https://www.cambridgecognition.com/products/cognitive-research/) measures executive functions, attention, psychomotor response speed, and social cognition.
3. A performance validity test assesses engagement and motivation (Green et al., 2003).
4. Performance evaluations: Participants rate their performance on each test using a 7-point Likert-scale.

Part 2: Experimental cognitive tasks and psychophysiology

After the break, participants complete brief computerised tasks and psychophysiological testing administered with E-Prime experimental software (https://pstnet.com/products/e-prime/) and a Powerlab psychophysiological data acquisition system (https://www.adinstruments.com/products/powerlab-daq-hardware). The measures are as follows:

1. Psychophysiology: Electrodermal activity (skin conductance) and heartrate (elctrocardiography) monitor autonomic activation.
2. Heartbeat tracking and time estimation tasks measure cardiac interoception and time perception respectively.
3. Computerised subjective probes assess momentary psychological and physical states (Table S1).
4. Grip contraction task: Participants repeatedly grip a dynamometer device with their dominant hand at maximum effort for short periods, separated by rests.

Participants are compensated with a £50 shopping voucher on completion of the laboratory session.

#### Neuroimaging

Participants eligible for MRI scanning return to complete the session on a separate day. The session lasts a maximum of 90-minutes, including the following components:

1. Structural scans (~10 mins).
2. Resting-state functional scan (~10 mins).
3. Interoceptive attention task: Participants direct their attention to their heartbeat sensations or an exteroceptive stimulus (~10 mins).
4. Affective images task: Participants view blocks of positive, negative and neutral images (International Affective Picture System; Lang et al., 2008). After each block, participants report on momentary physical and psychological states (Table S2). Heartrate and electrodermal activity are monitored throughout (~25-30 mins).
5. Interoceptive attention task repeated.

Participants are compensated with a £50 shopping voucher on completion of the session.

#### Remote monitoring phase (2-4 weeks duration)

The initial remote monitoring phase includes collection of data on participants’ behaviours, experiences and physiological signals for two weeks in their everyday lives.

Participants receive support and training from the research team on installation and use of the smartphone applications (Real Life Exp / Fitbit). The ecological momentary assessment (EMA) protocol involves brief electronic questions daily (6 times per day) on physical and psychological states, and daily events (Table S1). Each data collection point is triggered by a notification from the RealLife Exp application and takes 1-2 minutes to complete. The notifications are sent at pseudorandom intervals during waking hours (9am-9pm). The notifications do not arrive less than one-hour apart. Participants also have the option to enter data on salient daily events as they occur, at any time of the day or night. Two questions on subjective sleep quality (duration, disturbance) are administered each morning.

Participants are also asked to wear a FitBit Charge 5 wearable continuously to collect data on potential physiological triggers for FND symptoms, including:

- Sleep quality and duration – measured daily
- Activity – measured continuously
- Heartrate – measured continuously

After the initial two-week period, participants with FND and clinical controls are invited to participate in a further two weeks using the RealLife app to log salient daily events, whilst continuing to wear the Fitbit device.

All participants are compensated for completion of the remote monitoring phase with a digital £50 shopping voucher and are able to keep the Fitbit device for their personal use if they have demonstrated adequate engagement with the study (~80% EMA response rate).

#### Follow-up Procedures

Participants with FND and clinical controls are invited to complete 3-, 6- and 12-month follow-ups, which involve completing an abbreviated version of the original online questionnaire pack (Figure 1) and a brief call with a member of the research team. They are also invited to provide their Fitbit data for that follow-up period in pseudonymised format, which can be downloaded on request from the Fitbit consumer-facing platform. The research team are available to support the participant with all remote activities as needed.

Participants are compensated for each follow-up with a digital £10 shopping voucher.

#### End of Study Procedures

The end of the study is defined as the time at which the last participant completes the final research activity (remote follow-up). Following this, we will inform any regulatory bodies (i.e., the Research Ethics Committee) and the recruitment partners (FND charities and NHS Trusts). After completion of data analysis, the results will be disseminated at academic conferences and in peer-reviewed publications. We will share our findings with the FND-PCAP and other relevant stakeholders via social media, email lists, and/or press releases.

### Statistical considerations

#### Sample size

The overall target sample size was determined on the basis of a power calculation completed in G*Power (Faul et al., 2007). We estimated that for a mixed model ANOVA/ANCOVA, which will be used to test within- and between-groups variation on the primary endpoint of autonomic arousal, a total sample of 180 (n=45 in each 4 groups) would have 80% power to detect a small effect (f=0.1) with an alpha of 0.05. Therefore, a total sample of 200 participants entering the study will allow for up to five non-completing participants or missing data in each group.

On the basis of our pilot study data, we anticipate that approximately 90% of FND patients screened will be eligible and approximately 70% will proceed to complete the study; therefore, we plan to screen ~71 potential participants in each FND group (FS, FMS), to achieve the target sample size (FS n=50; FMS n=50). We predict that approximately 15 participants in each group will be ineligible, unsuitable, or unwilling to participate in the neuroimaging session; therefore, the anticipated sample size for this element of the project is n=35 in each of the four study groups (total n=140). For the remote monitoring data analysis, it is estimated that 45 participants per group will allow sufficient statistical power with four variables (affect, dissociation, HR, SCL) entered into a predictive model with momentary FND symptom ratings as the primary outcome variable (Peduzzi et al., 1996).

#### Data processing and statistical analyses

Detailed statistical analysis plans are under development for all elements of the project and these will be finalised and pre-registered before data analysis commences. The nature of all data types are outlined in Table S2.

The data will be checked for relevant assumptions (normality of distribution, sphericity, homogeneity of variances) prior to analysis. Non-normally distributed data will be transformed, or non-parametric or robust tests used. The amount and nature of missing data and outliers will be examined for all outcome variables. The approach taken with missing data will depend upon the number of missing values and whether the data is judged to be missing at random or missing not at random. The options for addressing missing data will include variable or participant exclusion, pairwise deletion, single or multiple imputation. Imputation methods will be used where missing data on a given measure does not exceed 20%. Outlying data points will be scrutinised and, where appropriate, the values will be Winsorized (replaced with less extreme values). The analyses will be repeated with the original and Winsorized values. Rates of missing data and outliers will be recorded and reported, along with any sensitivity analyses conducted.

Recruitment and retention rates will be assessed with descriptive statistics. Demographics and background variables will be analysed with appropriate between-group tests including t-tests, Mann-Whitney tests, and/or Analysis of Variance (ANOVA). Cognitive and experimental task data and autonomic measures will be analysed with between- or mixed between- and within-group tests, such as ANOVA/ANCOVA with covariates added where relevant, and/or multivariate regression analyses. Chi-squared tests will be used for categorical data.

Standardised data pre-processing and analysis methods will be adopted for analysis of neuroimaging data. Pre-processing will include reorientation, motion correction, spatial smoothing and normalisation. The neuroimaging data will be analysed with region-of-interest and network connectivity analyses. The primary regions of interest (ROIs) will be: amygdala, periaqueductal grey, and insula. Connectivity analyses will examine functional connectivity patterns between hubs in neural networks involved in affective activation and awareness (amygdala, periaqueductal grey, insula, anterior cingulate cortex) and those involved in motor and cognitive control (motor/premotor areas, dorsolateral prefrontal cortex).

The remote monitoring data will be analysed with multilevel modelling, to identify variables (e.g., dissociation, affect, autonomic arousal, life events) that are predictive of FND symptom occurrence/worsening in participants’ daily lives. Multilevel models are extensions of the traditional regression model, which account for the nested, hierarchical nature of intensive longitudinal data, by including random effects into model coefficients to account for within-participant variation over time. Furthermore, these techniques are suitable for use when there are missing data and unbalanced numbers of observations, as is often the case in remote measurement studies.

The alpha value to be adopted for most statistical tests will be p≤0.05; however, if multiple tests are conducted with related data (e.g., sub-scales of a questionnaire, post-hoc tests following ANOVA, exploratory tests of multiple neural regions [excluding ROIs]), we will use methods for controlling inflation of familywise error or false discovery rate, such as Bonferroni or Benjamini-Hochberg corrections respectively.

## Ethical considerations & regulatory approvals

The study is sponsored by King’s College London and approved by the North West – Greater Manchester South Research Ethics Committee (ref NW/23/0217). The research will be conducted in compliance with the principles of the Declaration of Helsinki (1996), and in accordance with all applicable regulatory requirements including but not limited to the UK policy framework for health and social care research, Trust and Research Office policies and procedures and any subsequent amendments. All participants will provide written informed consent prior to participation in the study, as described above (Methods section).

This protocol has been developed in consultation with the FND-PCAP and a team of academic and clinical collaborators with expertise in FND research and treatment, or relevant methodologies. The conduct, progress and results of the study will be reported to these groups, as well to relevant stakeholders (FND Hope UK, FND Action) and to the approving Research Ethics Committee and institutional research governance office (KCL).

## Risk management, adverse events & withdrawal

### Risk management & safeguarding

Some of the research activities could potentially be sensitive or challenging for some participants, such as answering questions regarding physical and mental health, difficult cognitive tests, exposure to emotionally significant stimuli, discomfort during the neuroimaging session, and multiple daily prompts during the initial remote monitoring phase. Our pilot study showed that most participants found these elements of the study acceptable/tolerable, although we have incorporated feedback to improve participants’ experiences.

The research team will ensure that participants are informed clearly about the nature of all tasks, before each activity commences. All participants will be warned that some of the activities might be challenging and that they may experience some degree of discomfort or stress during specific elements of the research. Participants will be reminded that they are free to decline to complete any aspect of the study, or that they may choose to decline specific measures or items, or withdraw from the study at any time, without explanation. The research team will be vigilant for signs of distress throughout the procedures and will pause or end procedures if deemed necessary to ensure the participant’s well-being. During remote monitoring, the research team will be available by telephone and email to answer questions, trouble-shoot and provide any reassurance that participants may require. The Chief Investigator (SP) will arrange to speak to a participant to discuss any distress experienced during the study, on request. A list of support organisations is included in the Participant Information Sheet as standard. Protocols are in place for safeguarding actions to be taken if a participant were to disclose a risk of harm to themselves or another, or if a severe and urgent mental health risk or existing harm became apparent during the course of the study. Adverse events occurring during the study will be monitored and recorded by members of the research team, followed-up by SP, and reported to the KCL Research Governance team and the Research Ethics Committee following established procedures.

### Withdrawal/ drop out

Premature withdrawal of participants may occur for the following reasons:

- participant no longer wishes / is able to take part;
- change in eligibility status is identified;
- a protocol violation.

If a participant withdraws, or is withdrawn from the study, the data collected from them up to that point will be retained in an anonymised format, and the reason for withdrawal will be recorded if voluntarily provided by the participant. No further data would be collected from participants who withdraw or who are withdrawn.

## Data management

All data are stored electronically in secure encrypted databases (e.g., MS Excel), on KCL SharePoint and OneDrive servers. Only immediate members of the research team have access to the databases, as needed. All research data are marked only with a Participant Identification Number (PIN), not with any identifiable data. The linkage data, in which the PIN and identifiable information are associated, is held on a password-protected database, securely on KCL servers. Participant identifiable data will not be shared with any third parties, nor will it be shared outside of European Union countries. Participants have the right to withdraw their identifiable data from the study.

The data that participants share with the research team via the GDPR-compliant Qualtrics, LifeData and Fitbit platforms does not contain any identifiable data and is extracted from the platforms by the research team as soon as possible and stored on secure KCL servers. Participants’ data will be deleted from the third-party platforms as soon as possible following data extraction. The data are anonymous to the third parties, who have no access to any linkage data, although the data are pseudonymous to the research team who will have access to linkage data.

In publications and other reports of the study findings, only aggregated group level data will be presented, in which no single individual would be identifiable. When the study is complete, the data will be held securely for ten years, as specified by the King’s College London data retention policy.

## Financing and Insurance

This study is funded by a Medical Research Council Career Development Award to SP [2022-2026, ref MR/V032771/1] and is part-funded by the National Institute for Health and Care Research (NIHR) Maudsley Biomedical Research Centre. The sponsor (KCL) will, at all times, maintain adequate insurance in relation to the study through its professional indemnity (Clinical Trials) & no-fault compensation in respect of any claims arising as a result of negligence by its employees, brought by or on behalf of a study participant.

## Publication and Dissemination

The anonymised, raw data set will be made available on reasonable request and with consultation with the KCL Data Management team. The research findings will be presented at academic conferences and published in peer-reviewed journal articles, as well as being disseminated to a wider audience through social media channels and/or other public relations and media outlets.

Participants will be informed that the results will be published and will be offered the opportunity to receive a summary of the publications and electronic links to the outputs on request.

## Discussion

Evidence for several hypothesised pathophysiological mechanisms in FND is limited at present due to methodological constraints in previous studies (Drane et al., 2020; Pick et a., 2019). This project employs a novel combination of multimodal research methods to rigorously examine several putative mechanisms in FND, at multiple levels of explanation, using subjective/experiential, behavioural, and physiological measures. The study is testing causal hypotheses about the role of altered emotional processing, autonomic arousal, dissociation and interoception in the initiation or exacerbation of FND symptoms, directly comparing these processes in FS and FMS. We aim to control, or account for, a range of common confounding factors, including comorbid psychiatric symptoms, medication use, and general cognitive functioning.

This is the first study to directly compare FND subgroups using such diverse and comprehensive methods in a relatively large sample, including both clinical and healthy comparison groups. By examining these mechanistic processes in both laboratory and naturalistic contexts, we will ascertain the consistent influences and underlying processes that might explain the occurrence of FND symptoms across settings. The combination of neuroimaging, psychophysiology, and momentary subjective probes will allow us to examine the simultaneous neural, autonomic, and psychological processes that precede subtle variations in subjective FND symptoms from moment-to-moment. The use of remote monitoring technologies will highlight the physiological, psychological and environmental factors influencing FND symptoms in real-time and real-world settings. Moreover, the inclusion of longitudinal follow-ups will establish whether specific aetiological factors and mechanistic features are predictive of longer-term clinical outcomes in FS and FMS.

The results of this study have the potential to lead to significant progression in aetiological and mechanistic models of FND, and will likely uncover meaningful, evidence-based targets for prevention and intervention of FND in future.

## Funding

The study is funded by a Medical Research Council Career Development Award to SP [MR/V032771/1]. This project also represents independent research part-funded by the National Institute for Health and Care Research (NIHR) Biomedical Research Centre at South London and Maudsley NHS Foundation Trust and King’s College London. The views expressed are those of the authors and not necessarily those of the NHS, the NIHR or the Department of Health and Social Care.

## Conflicts of interest

None.

## Author contributions

Funding acquisition: SP (lead), MH, TC, MAM (supporting). Conceptualisation: SP. Design/methodology: SP (lead), MH, TC, MAM, ASD, LSMM, AATSR, MJE, LHG, TRN, JSW (supporting). Project administration: SP. Resources: SP (lead), MH, TC (supporting). Writing: SP (original draft/review and editing), MAM, TC, MH, LSMM, LHG, MJE, TRN, JSW, ASD, BS (review and editing). All authors reviewed and approved the final version of the manuscript.

## Data Availability

Participant identifiable data will not be shared with any third parties, nor will it be shared outside of European Union countries. Anonymised raw data will be made available on reasonable request, in consultation with the King's College London data management team, and after suitable data sharing agreements have been completed.

## Notes

### Competing Interest Statement

The authors have declared no competing interest.

### Author Declarations

The study is sponsored by King’s College London and approved by the North West – Greater Manchester South Research Ethics Committee (ref NW/23/0217). The research will be conducted in compliance with the principles of the Declaration of Helsinki (1996).

